# Measurement of small droplet aerosol concentrations in public spaces using handheld particle counters

**DOI:** 10.1101/2020.10.13.20211839

**Authors:** G. Aernout Somsen, Cees van Rijn, Stefan Kooij, Reinout A. Bem, Daniel Bonn

**Author notes:** Correspondence: Prof. Dr. Daniel Bonn, Institute of Physics, University of Amsterdam, Science Park 904, 1098XH Amsterdam, The Netherlands.

## Abstract

We investigate the role of aerosols in the transmission of SARS-CoV-2 in public spaces. Direct measurement of aerosol concentrations however has proven technically difficult; we propose the use of handheld particle counters as a novel and easily applicable method to measure aerosol concentrations. This allow us to perform measurements in typical public spaces, each differing in volume, number of people and ventilation rate. These data are used to estimate the relation between aerosol persistence time and the risk of infection with SARS-CoV-2.

## Introduction

The World Health Organization has in its recent Scientific Brief^1^ highlighted the possible role of aerosols in the transmission of SARS-CoV-2^1-4^ and stated that ‘much more research is needed given the possible implications of such route of transmission’ ^1^. This is particularly relevant for public spaces where the risk of aerosol transmission of SARS-CoV-2 is highest. Direct measurement of aerosol concentrations however has proven technically difficult^5^, hampering such research. We validate the use of handheld particle counters as a novel and easily applicable method to measure aerosol concentrations.

To demonstrate the usefulness of our novel method, we perform measurements in typical public spaces that can play a role in aerosol transmission of SARS-CoV-2, each differing in volume, number of people and ventilation rate. These data are used to estimate the relation between aerosol persistence time and the risk of infection with SARS-CoV-2.

## Methods

Aerosol concentration is often measured using a laser sheet diffraction technique, in which the number of pixels that light up is a measure for the number and volume of the droplets.^4^ However, this technique can only be operated by highly specialized personnel and, because of laser safety issues, only in laboratory settings. Using this technique as the standard, we validate a novel method using a handheld particle counter (Fluke 985, Fluke B.V. Europe, Eindhoven, The Netherlands) which is frequently used for air quality assessment and overcomes most of the above-mentioned drawbacks of the laser sheet diffraction technique. For each measurement, the aerosol concentration is determined by correcting for the background (measured for ∼8 minutes), consisting of dust particles.

### Validation

First, we validate the novel method by comparing the results with those obtained previously using the laser diffraction technique^4^. The validation scenario is that of a single person coughing once inside a poorly ventilated restroom of volume 8 m^3^. The results for both techniques are shown in Fig. 1. We find that coughing leads to the generation of an amount of aerosol particles an order of magnitude above the background level of the particle counter; both techniques next reveal that the number of aerosols per liter of air decreases exponentially in time, with a time constant of ∼4 minutes (Fig. 1). Fig. 2 shows the size distribution of the particles as obtained by the two techniques. The comparison is more difficult here, as the handheld particle counter has less channels to separate ranges of particles sizes as the laser sheet diffraction technique; however, the results are not incompatible. We also find that the results obtained using the handheld particle counter were reproducible to within 10%. We conclude that the particle counter technique is indeed a reliable method to determine aerosol concentrations as a function of time, as well to give a rough indication of the size distribution of droplets.

**Figure 1:**
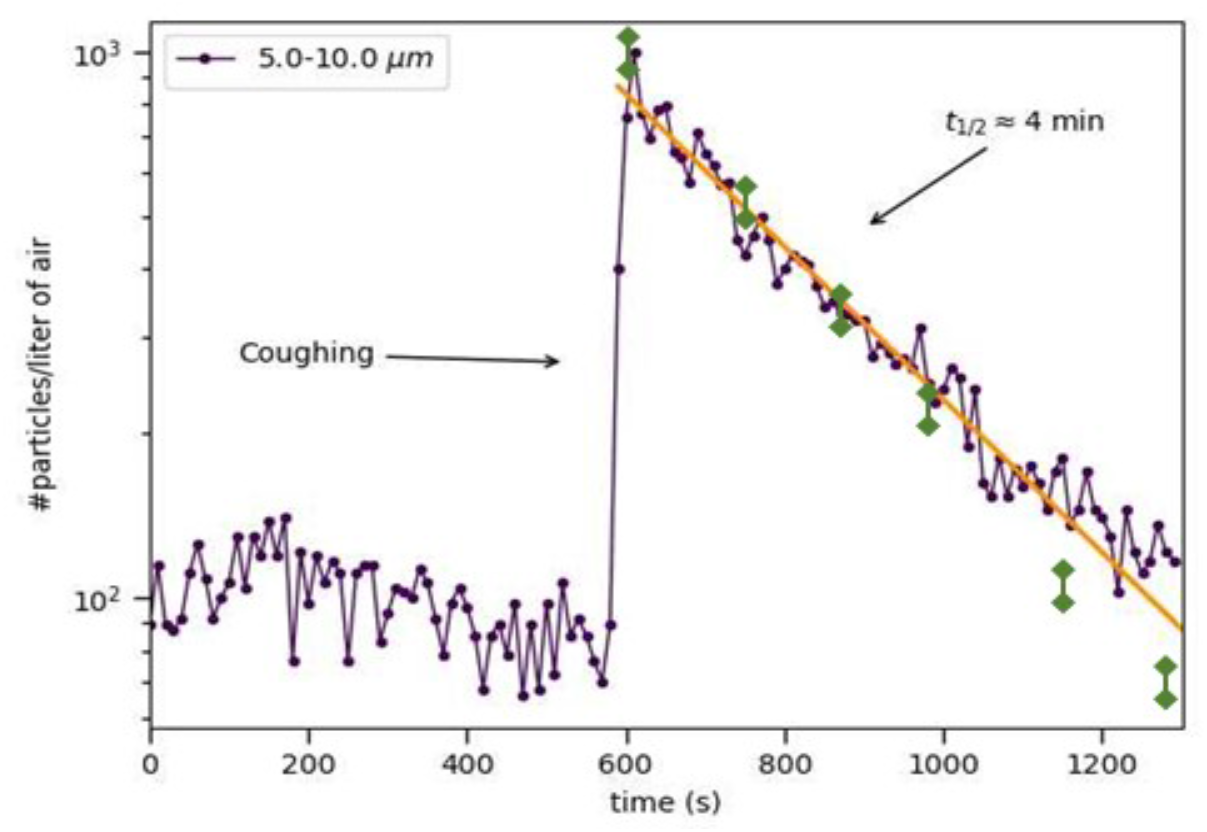
Concentration of airborne particles of diameters 5.0-10.0 µm as a function of time as measured using a handheld particle counter (purple symbols) and the laser sheet diffraction technique^4^ in a poorly ventilated space of volume 8 m^3^. The arrow indicates the moment of coughing, coinciding with a sharp increase followed by an exponential decay, with a half-life of roughly 4 min. The yellow line is a guide to the eye. The green data points are the reference data from the laser sheet technique published previously in [4]

**Figure 2:**
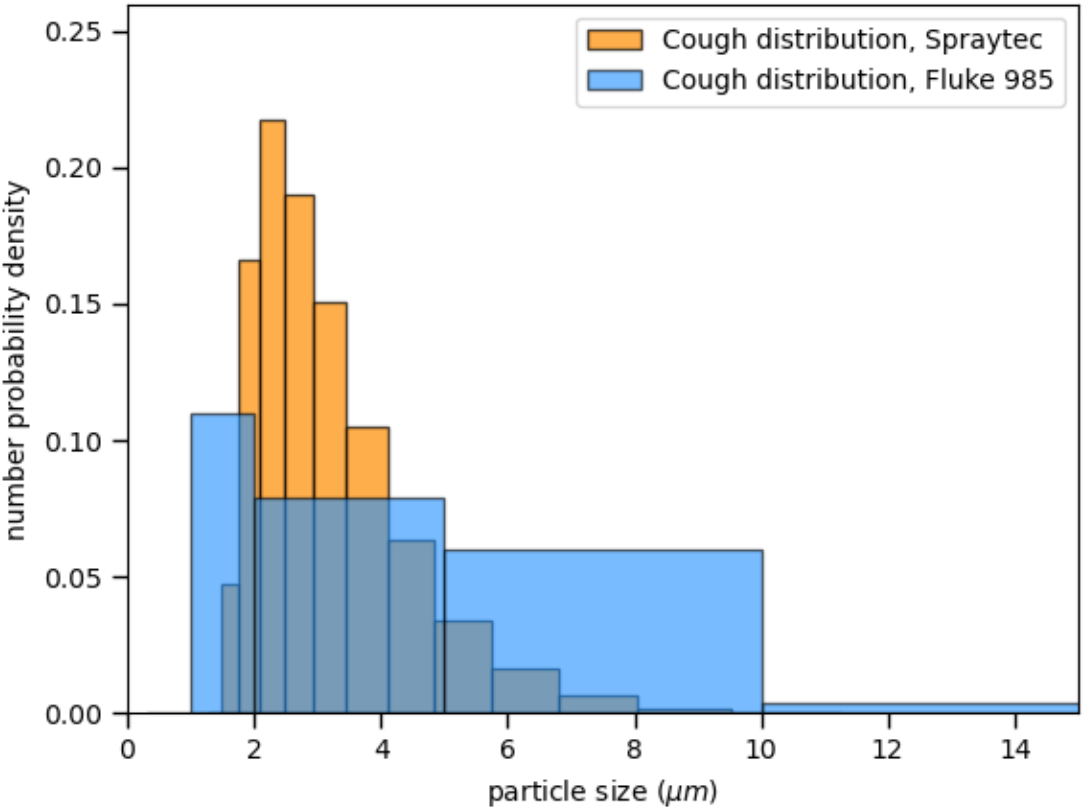
Number particle size distributions measured by laser diffraction (yellow bars) compared with the distribution inferred from the particle counter (blue bars). The limited number of channels of the particle counter makes for a crude distribution, but the two are compatible.

### Application to public spaces

We now use the technique to characterize a wide range of real-world public spaces, each selected as a typical example of a certain type of public space (Table 1). In these public spaces, we measure all aerosols resulting from breathing, speaking, coughing and sneezing by people present (1 to 25 at the time of the measurements). Background measurements indicate that dust generated by the people moving around is mostly (>98%) contained in the first two channels of the particle counter (diameters between 0.3 and 0.5 micrometer), out of the range of the characteristic diameter of aerosols produced by breathing, speaking, coughing and sneezing^4^. They are therefore not considered when evaluating the aerosol density. Separate from the measurements involving multiple people producing aerosols, we also investigate the decrease in aerosol concentration over time by generating a known quantity of artificial aerosols using a specially designed spray nozzle as used in [4], which is known to produce aerosols of the same size distribution as respiratory droplets resulting from coughing (i.e., between 1-10 micrometer with a maximum at 4 micrometer). In Table 1 we summarize the results, indicating the number of people involved, the air change rate per hour (ACH) of the ventilation system used, and the measured droplet concentration half-times.

**Table 1:** Aerosol concentrations and persistence times in different public spaces characterized by the number of people present, volume and rate of ventilation. In all but the club scenario where aerosols where artificially generated, the aerosol origin is from persons speaking, coughing and sneezing. Spaces were sampled that gave us permission to do so-these were mainly well-ventilated spaces in modern buildings. Each space was typically sampled in 5 different places.

| SPACE | SIZE (M<sup>3</sup>) | AIR CHANGE PER HOUR (H<sup>-1</sup>) | AEROSOL ORIGIN | 50% DECREASE (MIN) | AEROSOL PART/L | RNA COPIES/L | COVID-19 INFECTION RISK |
| --- | --- | --- | --- | --- | --- | --- | --- |
| GYM | 2000 | 5-15 | 25 visitors | 1 | <10 | <1 | Low |
| TRAIN | 150 | 0-5 | 20 visitors | 2 | 210 | <21 | Low |
| MEETING ROOM | 30 | 10 | 4 visitors | 1 | 45 | <5 | Low |
| NIGHT CLUB | 2000 | 5-15 | Artificial | 1 | <10 | <1 | Low |
| CAR | 3 | 5-20 | 2 visitors | 0.5 | 20 | <2 | Low |
| AIRPORT | 12000 | 5-15 | ~100 visitors | 1 | <10 | <1 | Low |
| RESTAURANT | 120 | 8 | 25 visitors | 1 | 248 | <25 | Low |
| RESTROOM | 8 | ~1 | 1 visitor | 4 | 7716 | <772 | Intermediate |
| OFFICE SPACE | 50 | 10 | 5 visitors | 1 | 35 | <4 | Low |
| LUNVENTILATED LIVING ROOM | 80 | ~1 | 4 visitors | 5 | 5214 | <520 | Intermediate |
| ELEVATOR | 8 | ~1-4 | 2 visitors | 5 | 4350 | <435 | Intermediate |

In the public spaces investigated by us, aerosol concentrations are approximately 20 to more than a 100 times lower in all ventilated public spaces compared to the poorly ventilated restroom used for the calibration measurements. Also in a public elevator and in a poorly ventilated living room, aerosol concentrations are high. The characteristic times for a 50% decrease in aerosol concentration are on the order of one minute in well-ventilated spaces, compared to 4-5 minutes in the poorly ventilated restroom, elevator and living room.. This is due to both the air renewal (given by the ACH) and further dilution by dispersion throughout the space, and therefore depends both on the ACH and the absolute size of the given space (Table 1).

## Discussion

This study shows that the particle counter technique is a reliable method to investigate aerosols concentrations and their evolution in time. Using this easily applicable method, aerosol concentrations can be measured in any public space which is important to determine the risk of aerosol transmission of SARS-CoV-2 and to evaluate the impact of risk reducing measures (i.e. improving ventilation).

Aerosol persistence times in the tested spaces are relatively short due to adequate space ventilation. Current standards for the air change rate by mechanical room ventilation vary between 4-20 air changes per hour. An air change rate of 10 times per hour means that every 6 minutes the given space has received fresh air of a volume similar to that of the spaceOur measurements of aerosol persistence suggest that a half-life of the aerosol concentration of one or two minutes minimizes the aerosol concentration, and is achieved for air change rates in excess of 10 air changes/hour. Using the half-times as measured by us, we calculate that the decrease in the number of aerosol particles after these 6 minutes will vary between 50 and 100%, depending on the ventilation method and the size of the public space.

When translating our findings to practical risk assessments in the context of SARS-CoV-2 transmission by aerosols, we conclude that the risk of transmission via airborne aerosols is low in all but the restroom scenario. The reason for this is twofold: first, good ventilation significantly decreases the density of aerosols in a short time (Table 1). Second, the number of viral particles in the very small aerosol drops is low. Sputum droplets from COVID-19 patients carry typically between 10^4^ -10^9^ RNA copies per ml^5^. This implies between 0.001 and 100 RNA copies per thousand aerosol drops^6,7^. The minimum infectious dose for SARS-CoV-2 has not been reported; the severity of COVID-19 is however believed to be proportional to the dose of the initial inoculum, implying that transmission by aerosols may lead to relatively milder symptoms^6,7^. In our risk analysis we assume that exposure to less than 10^3^ microdroplets (corresponding to less than 100 RNA copies) imposes a low risk, between 10^3^ and 10^5^ microdroplets (corresponding to 100 - 10.000 RNA copies) an intermediate risk, and more than 10^5^ micro droplets (corresponding to more than 10.000 RNA copies) a high risk of transmission^6,7^. Further research on the transmissibility of the virus is needed to validate this assumption and possibly correct these risk values. Our measurements together with the risk analysis however underlines the importance of good ventilation, and suggests that health authorities can advise a minimal ventilation rate to minimize the probability of SARS-CoV-2 transmission in public spaces.

## Conclusion

This study demonstrates a novel aerosol measurement method that is easily implemented in different environments. With reasonable assumptions on the viral load and infectivity, this allows for a rough estimate of the probability of aerosol transmission of SARS-CoV-2 following [6,7]. To reduce the spread of such infections, healthcare authorities should consider this method to evaluate the ventilation of public spaces, especially in spaces where aerosolization is common such as hospital and dentistry settings.

## Data Availability

All data are available from the corresponding author upon reasonable request

